# Effectiveness of the BNT162b vaccine fourth dose in reducing SARS-CoV-2 infection among healthcare workers in Israel, a multi-center cohort study

**DOI:** 10.1101/2022.04.11.22273327

**Authors:** Matan J Cohen, Yonatan Oster, Allon E Moses, Avishay Spitzer, Shmuel Benenson, the Israeli-hospitals 4^th^ vaccine Working Group

## Abstract

During December 2021 the fifth COVID-19 wave started in Israel, caused mostly by the Omicron variant, affecting the unvaccinated and vaccinated population. Ninety percent of the Israeli adults, including most healthcare workers (HCWs), received three doses of the BNT162b2 vaccine until September 2021. Following the success and safety of the 3^rd^ dose in preventing infection and severe disease, on December 30, 2021, the Israeli Ministry of Health recommended a voluntary 4^th^ vaccine dose to adults above 60 years, immunocompromised, and HCWs. We compared breakthrough infections in HCWs, between 3 and 4-dose recipients.

Hospitals collected data on personnel vaccinations and infections dates. The study cohort included all HCWs in eleven hospitals in Israel, who have been vaccinated with three doses up to September 30, 2021, and had not contracted COVID-19 before the vaccination campaign (January 2, 2022).

We calculated breakthrough infection rates in 4-dose recipients (more than six days after vaccination) vs. 3-dose recipients. Rate-ratios were calculated for the entire cohort and for subgroups (hospital, sex, age-groups, and profession). Additionally, we repeated the calculations on 4-dose and 3-dose recipients who received the 3^rd^ dose on the same date and were matched for sex, age group, profession and hospital. We generated time-dependent Cox-regression models to account for 4^th^ dose administration timing (Supplement).

There were 29,612 HCWs who received 3 vaccine doses between August and September 2021; of these, 5,331 (18.0%) received the 4^th^ dose during January 2022 and were not infected by the first week after vaccination. Overall breakthrough infection rates in the 4-dose and 3-dose groups were 368/5331 (6.9%) and 4802/24280 (19.8%), respectively. The RR (95%CI) was 0.35 (0.32 to 0.39) for crude analysis, and 0.61 (0.54 to 0.71) in the matched analysis. The adjusted HR in the Cox-regression model was 0.56 (0.50 to 0.63). In both groups, severe disease and death were not reported.

Our data shows that the 4^th^ BNT162b2 dose resulted in reduced breakthrough infection rates among HCWs. This reduction, similar to the findings in the Israeli elderly population, is lower than that observed after the 3^rd^ dose.

Nevertheless, considering the high infectivity of the Omicron variant, which led to critical medical staff shortages, a 4^th^ vaccine dose should be considered to mitigate the infection rate among HCWs.

## Introduction

During December 2021 the fifth COVID-19 wave started in Israel, caused mostly by the Omicron variant, affecting the unvaccinated and vaccinated population (1, 2). Ninety percent of the Israeli adults, including more than 95% of healthcare workers (HCWs) (3), received three doses of the BNT162b2 vaccine until September 2021. Following the success and safety of the 3^rd^ dose in preventing infection and severe disease (4), and assuming waning immunity of the 3^rd^ dose, the Israeli Ministry of Health recommended a voluntary 4^th^ BNT162b2 dose to adults above 60 years, immunocompromised, and HCWs. We compared breakthrough infections in HCWs, between 3 and 4-dose recipients.

## Methods

Hospitals collected data on personnel vaccinations and infections dates. The study cohort included all HCWs in eleven hospitals in Israel, who have been vaccinated with three doses up to September 30, 2021, and had not contracted COVID-19 before the vaccination campaign (January 2, 2022). The participating centers included about a half of the total acute care beds in Israel.

We calculated breakthrough infection rates in 4-dose recipients (at least one week after vaccination) vs. 3-dose recipients. Workers were tested by PCR at their own decision; there was no systematic testing. Rate-ratios were calculated for the entire cohort and for subgroups (hospital, sex, age-groups, and profession). Additionally, we performed calculations on 4-dose and 3-dose recipients who received the third dose on the same date and were matched for sex, age group, profession and hospital. We generated time-dependent Cox-regression models to account for fourth dose administration timing (Supplement). The study was approved by the local ethics committee of each center:

a. Reviewed by the ethical review board, Bnei Zion Medical Center, Haifa, Israel, and ethical approval was given, need for consent was waived.
b. Reviewed by the ethical review board, Tel Aviv Sourasky Medical Center, Tel Aviv, Israel, and ethical approval was given, need for consent was waived.
c. Reviewed by the ethical review board, Edith Wolfson Medical Center, Holon, Israel, and ethical approval was given, need for consent was waived.
d. Reviewed by the ethical review board, Shaare-Zedek Medical Center, Jerusalem, Israel, and ethical approval was given, need for consent was waived.
e. Reviewed by the ethical review board, Hadassah-Hebrew University Medical Center, Jerusalem, Israel, and ethical approval was given, need for consent was waived.
f. Reviewed by the ethical review board, Meir Medical Center, Kfar-Saba, Israel, and ethical approval was given, need for consent was waived.
g. Reviewed by the ethical review board, Kaplan Medical Center, Clalit Health Services, Israel, and ethical approval was given, need for consent was waived.
h. Reviewed by the ethical review board, Rambam Health Care Campus, Haifa, Israel, and ethical approval was given, need for consent was waived.
i. Reviewed by the ethical review board, Shamir (Assaf Harofeh) Medical Center, Zerifin, Israel, and ethical approval was given, need for consent was waived.
j. Reviewed by the ethical review board, Baruch Padeh Medical Center, Poriya, Israel, and ethical approval was given, need for consent was waived.
k. Reviewed by the ethical review board, Barzilai Medical Center, Ashkelon, Israel, and ethical approval was given, need for consent was waived.

## Results

There were 29,612 HCWs who received 3 vaccine doses between August and September 2021; of these, 5,331 (18.0%) received the 4^th^ dose during January 2022 and were not infected by the first week after vaccination (supplement Figure). Factors linked to higher vaccination rate were male sex, older age and medical profession (supplement Table 1).

HCWs characteristics and 4-dose vaccination rates were similar in all participating hospitals (supplement Table 2). Overall breakthrough infection rates in the 4-dose and 3-dose groups were 368/5331 (6.9%) and 4802/24280 (19.8%), respectively. The RR (95%CI) was 0.35 (0.32 to 0.39) for crude analysis, and 0.61 (0.54 to 0.71) in the matched analysis. The adjusted HR in the Cox-regression model was 0.56 (0.50 to 0.63). Overall, the effect of the 4^th^ dose on COVID-19 infection rate was consistent over crude, matched and modelled analyses, in all subgroups (Table). Kaplan-Meyer curves of the crude and matched data are presented in the Figure. In both groups, there were no severe disease and death.

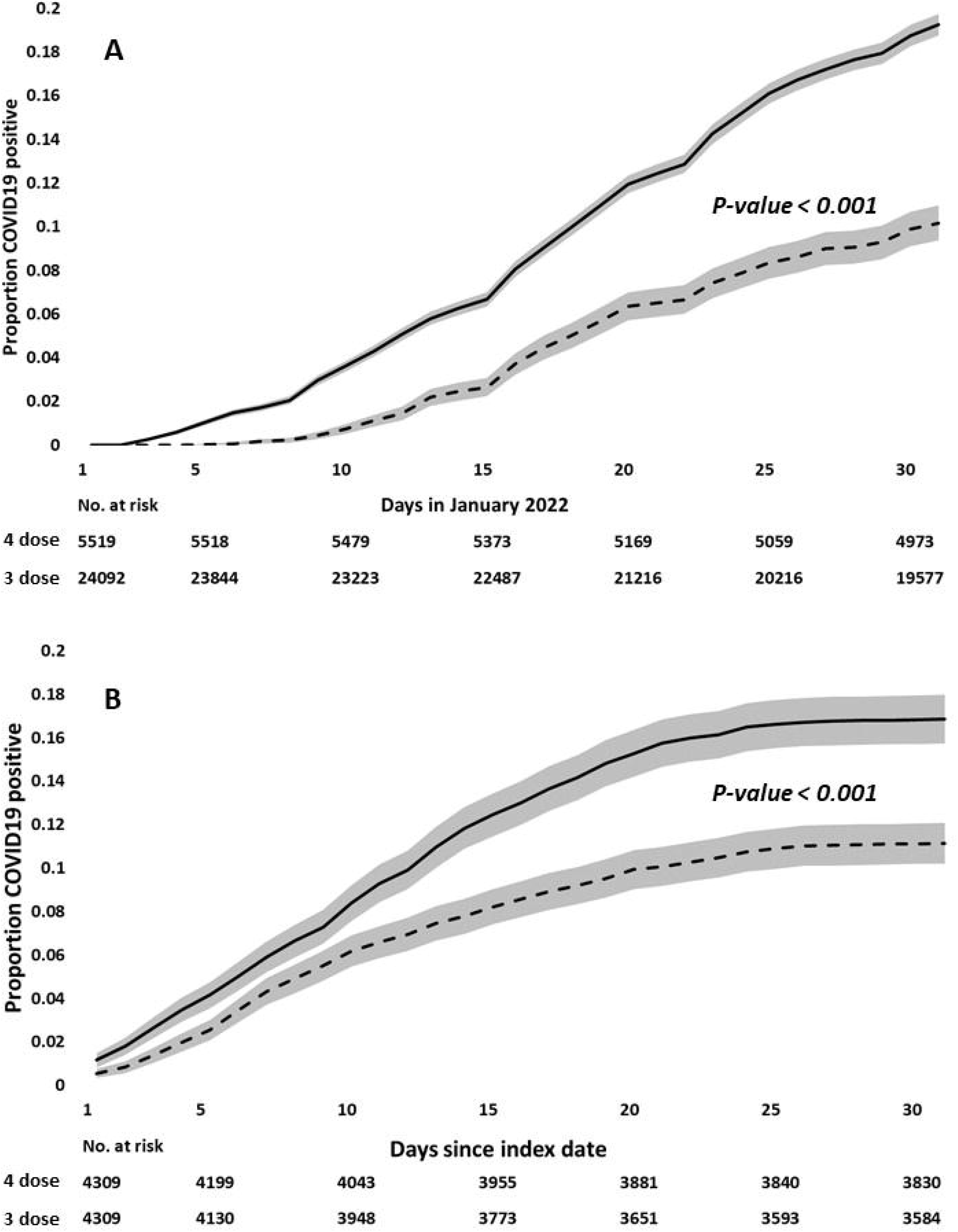
Morbidity curves of the cumulative COVID-19 rate estimate and confidence intervals (grey shadings) among healthcare workers who received four vaccine doses (dashed line) and three vaccine doses (continuous line). A – Proportion of breakthrough COVID-19 infections in the entire study cohort during January 2022 (*p-value* < 0.001). B - Proportion of breakthrough COVID-19 infections in the matched study cohort. For each matched pair, the index date was the day the worker received the 4^th^ vaccine dose (*p-value* < 0.001).

## Conclusions

Our data shows that the 4^th^ BNT162b2 dose resulted in reduced breakthrough infection rates among HCWs in 11 hospitals. This reduction is similar to the findings in the elderly Israeli population (5).

Though less effective than the 3^rd^ dose (6), this reduction along with a high HCWs infection rate results in a number-needed-to-vaccinate to prevent one infection of 7 to 13. Full vaccination would have decrease HCWs infection rate from 20% to between 6% and 12%.

Our study has some limitations. First, HCWs were not tested on routine basis; therefore, some infections might have been missed. Secondly, since the decision to receive the 4^th^ vaccine was on a voluntary basis, it is possible that those who chose to receive the 4^th^ dose were more careful to avoid getting infected then the others.

In conclusion, due to the high infectivity of the Omicron variant, and the demonstrated 4^th^ dose effectiveness, this booster should be considered for lowering the HCWs infection rate and critical staff shortages.

## Data Availability

All data produced in the present study are available upon reasonable request to the authors

## References

1. Israeli Ministry of Health COVID-19 data center. Accessed February 21, 2022. https://data.gov.il/dataset/covid-19 (In Hebrew).

2. Nemet I, Kliker L, Lustig Y, et al. Third BNT162b2 Vaccination Neutralization of SARS-CoV-2 Omicron Infection. N Engl J Med. 2022;386(5):492–494. doi:10.1056/NEJMc2119358

3. Patalon T, Gazit S, Pitzer VE, Prunas O, Warren JL, Weinberger DM. Odds of Testing Positive for SARS-CoV-2 Following Receipt of 3 vs 2 Doses of the BNT162b2 mRNA Vaccine. JAMA Intern Med. 2022; 182(2):179–184. doi:10.1001/jamainternmed.2021.7382

4. Bar-On YM, Goldberg Y, Mandel M, et al. Protection of BNT162b2 Vaccine Booster against Covid-19 in Israel. N Engl J Med. 2021;385(15):1393–1400. doi:10.1056/NEJMoa2114255

5. Bar-On YM, Goldberg Y, Mandel M, et al. Protection by 4th dose of BNT162b2 against Omicron in Israel. Medrxiv 2022.02.01.22270232; doi: https://doi.org/10.1101/2022.02.01.22270232

6. Oster Y, Benenson S, Nir-Paz R, Buda I, Cohen MJ. The effect of a third BNT162b2 vaccine on breakthrough infections in healthcare workers: a cohort analysis [published online ahead of print, 2022 Feb 7]. Clin Microbiol Infect. 2022; S1198-743X (22)00043-X. doi:10.1016/j.cmi.2022.01.019

