## Supplementary for "Effectiveness of the BNT162b vaccine fourth dose in reducing SARS-CoV-2 infection among healthcare workers in Israel, a multi-center cohort study"

1. Introduction
2. Methods
3. Figure
4. Tables
5. References

**Introduction**

The COVID-19 Omicron wave in Israel started in December 2021, resulting in the highest infection rate of all previous waves (1). Israel was the first country to administer a third vaccine dose to the entire adult population, beginning in August 2021; 90% of the adult population was vaccinated, including more than 95% of healthcare workers (HCWs) (2). This third dose (booster) vaccine dramatically reduced the rate of breakthrough infections, severe disease and mortality during the Delta variant wave (3). Despite this high vaccination rate, the highly infectious Omicron variant caused a significant number of breakthrough infections among the thrice vaccinated. This led to the precedent decision of the Israeli Ministry of Health to recommend a 4^th^ vaccine dose to the adult population for those older than 60 years, the immunocompromised and to HCWs, to overcome the presumed declined immunity of vaccinated individuals. We examined the effectiveness of this fourth dose in reducing the rate of breakthrough infections among HCWs in several Israeli hospitals, during the peak of Omicron activity.

**Methods**

*Settings*

Eleven Israeli hospitals participated in the initial study: all are academic centers, five of them tertiary care centers. All HCWs were vaccinated by Pfizer's BNT162b2 mRNA vaccines. The study was approved by the local ethics committee of each center. In total, the participating centers included about a half of the total acute care beds in Israel and cover all areas of Israel.

*Data collection*

The study cohort included all HCWs who have been vaccinated with three BNT162b2 doses and had not contracted COVID-19 before January 3, 2022. We collected anonymized personnel demographics (sex, age group, profession) and vaccination and breakthrough infection dates for all participants, until January 31^st^, 2022. 98.9% of 4-dose recipients received their 3^rd^ dose during August and September 2021. We compared the sex, age and profession of those who were vaccinated during these months to the entire cohort, with similar distributions (Data not shown). Therefore, we limited the analysis and comparison of 3-dose and 4-dose recipients to the HCWs who were vaccinated with the third dose only during August and September 2021.

We compared the relative risk and hazard ratio (as detailed below) of breakthrough COVID-19 infections between HCWs who received three and four vaccine doses. In unmatched analyses, these measures of association were compared between 4-dose recipients who were not infected at least a week after receiving the 4^th^ dose and workers who received only three doses (this group included 188 4-dose recipients, who had contracted COVID-19 within a week from the fourth dose, before the protecting effect of this vaccine). Analyses were done for the entire cohort and stratified per subgroups: hospital, sex, age group (under 40 years, 40 to 59, 60 and older) and profession (physician, nursing, other). We generated an inverse Kaplan-Meier survival curve demonstrating the proportion of HCWs who contracted COVID-19 in each group (three and four vaccine doses groups), during January 2022. Curves were compared with the log-rank test.

We performed two additional analyses to assess the robustness of the results: 1) Matched analysis – we matched HCWs who received the 4^th^ dose to those who received only three doses (1:1 matching). For each 4-dose vaccinated participant a 3-dose vaccinated participant was matched by sex, age group, profession, hospital and date of receiving the third vaccine dose (exact matching). Matching was done only if the 3-doses participant was not infected by the date the matched 4-dose partner was vaccinated. If multiple potential matching partners existed, one was randomly selected. We used the McNemar test for the matched analysis; confidence intervals for the relative risk were calculated using the Wald method. We then generated a similar inverse Kaplan-Meier survival curve for the matched analysis. We used the Prentice-Wilcoxon test and the Gehan test for comparison of paired data. These analyses were performed using the WINPEPI software (Version 11.65).

2) Regression modelling - the timing of receiving the 4^th^ dose was at the participants’ discretion and could have taken place at any day after recommending the 4^th^ vaccine for HCWs and therefore, exposure status to the 4^th^ dose varied over time. To account for the changing exposure status throughout the study period, a Cox-regression analysis with exposure status as a time-varying covariate was conducted, evaluating the hazard-ratio associated with 4^th^ dose exposure status among HCWs who received three vaccine doses during August and September 2021. All participants were defined as unexposed at study initiation and as exposed seven days after receiving the 4^th^ dose. Participants were censored due to a confirmed SARS-CoV-2 infection or at the end of the study period. The Cox-regression model was adjusted for sex, age group, profession, hospital and month of receiving of 3^rd^ vaccine dose. All covariates were tested with Schoenfeld residuals to evaluate the assumption of hazard proportionality. Non-proportional hazards were handled using stratification. This unmatched model was followed with a model including complete matching as described in the previous analysis.

Additionally, we planned to calculate the *number-needed-to-vaccinate* to prevent a single COVID-19 infection in a HCW by multiplying the calculated relative-risk or hazard-ratio by the base rate of COVID-19 in the 3-dose group. This results in the expected COVID-19 rate in the 4-dose group. Dividing 1 by the rate difference results in the *number-needed-to-vaccinate.*

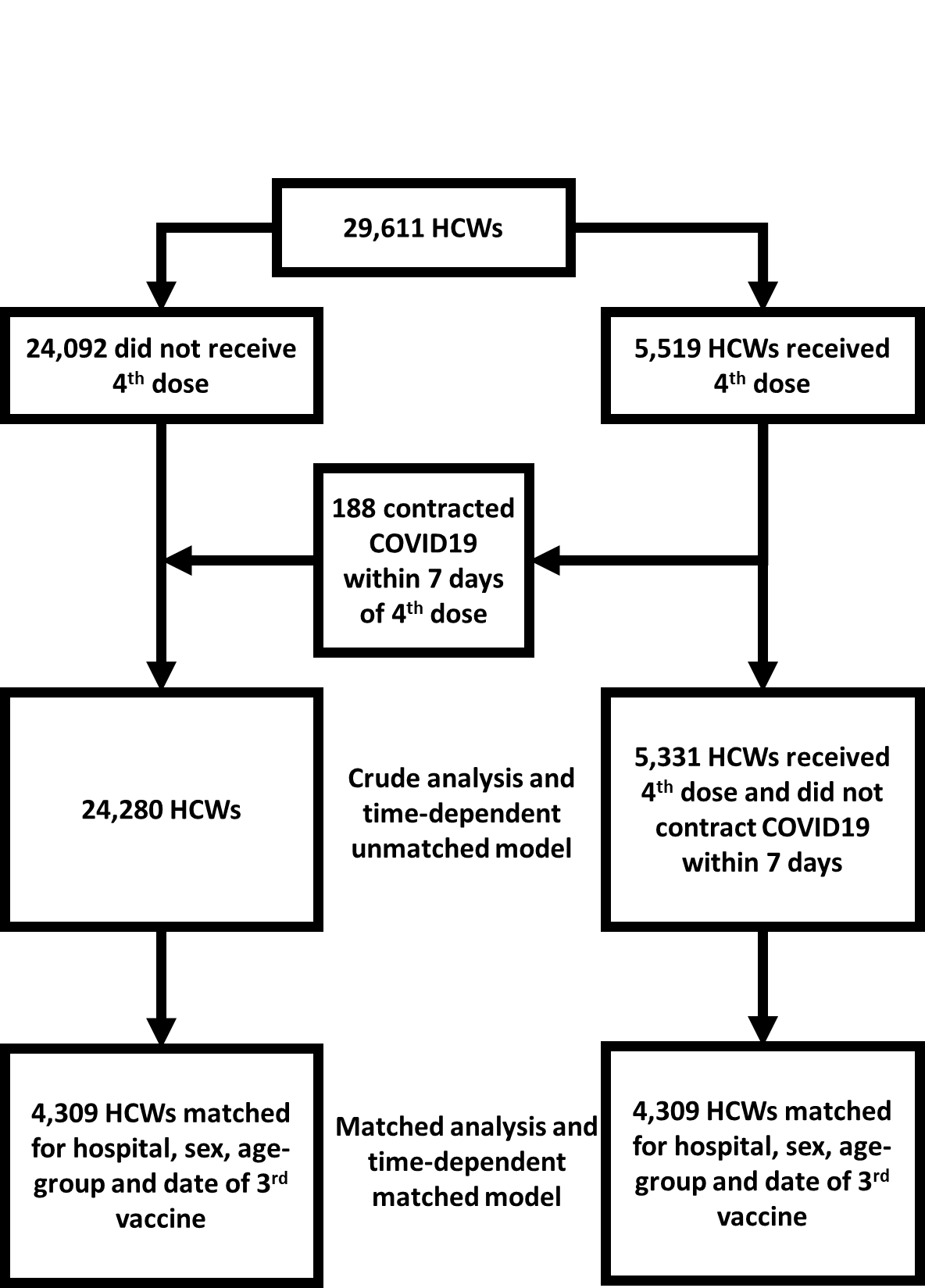

**Figure:** Flow chart of the analyses of breakthrough infections among 3 and 4 vaccine dose recipients, among healthcare workers in nine hospitals in Israel.

| **Supplement Table 1:** Characteristics of 29,612 healthcare workers from eleven hospitals in Israel, by number of vaccine doses* | | |
| --- | --- | --- |
|  | 3-dose group | 4-dose group |
| **Number of workers** | **24092** | **5519** |
|  | N (%) | |
| **Sex** |  |  |
| Female | 16365 (84%) | 3016 (16%) |
| Male | 7727 (75%) | 2503 (25%) |
| **Age group (years)** |  |  |
| Under 40 | 10376 (90%) | 1165 (10%) |
| 40 to 59 | 11293 (81%) | 2605 (19%) |
| 60 and older | 2423 (58%) | 1749 (42%) |
| **Profession** |  |  |
| Physician | 5483 (74%) | 1887 (26%) |
| Nursing | 7731 (86%) | 1215 (14%) |
| Other | 10878 (82%) | 2417 (18%) |
| * 188 HCWs who were vaccinate with the 4^th^ dose contracted COVID-19 within 1 week from vaccination and were considered therefore included in the 3-dose group in the analysis. | | |

| **Supplement Table** **2**: Number of participating healthcare workers, by hospital and profession; number of workers which received 3 or 4 doses of the BNT162b2 vaccine | | | | | | | | | | | | |
| --- | --- | --- | --- | --- | --- | --- | --- | --- | --- | --- | --- | --- |
|  | BMC | BPMCP | BZMC | HHUMC | KMC | MMC | RHCC | SMC | SZMC | TASMC | WMC | **Total** |
| **N*** | 1382 | 1215 | 1607 | 3900 | 1872 | 2581 | 4457 | 2451 | 2864 | 5595 | 1723 | **29611** |
|  | N (%) | | | | | | | | | | | |
| **Profession** | | | | | | | | | | | | |
| Physician | 351(25) | 321(26) | 382(24) | 807(21) | 470(25) | 623(24) | 1022(23) | 618(26) | 827(29) | 1467(26) | 482(28) | 7370 (25) |
| Nursing | 522(38) | 369(30) | 477(30) | 1167(30) | 690(37) | 955(37) | 1051(24) | 857(35) | 1005(35) | 1241(22) | 612(36) | 8946 (30) |
| Other | 509(37) | 525(43) | 748(47) | 1926(49) | 712(38) | 1003(39) | 2384(53) | 940(39) | 1032(36) | 2887(52) | 629(37) | 13296 (45) |
| **Vaccine doses** | | | | | | | | | | | | |
| 3 Doses | 1228(89) | 1100(91) | 1282(80) | 3038(78) | 1508(81) | 2019(78) | 3787(85) | 2070(86) | 2166(76) | 4362(78) | 1532(89) | 24092(81) |
| 4 Doses | 154(11) | 115(9) | 325(20) | 862(22) | 364(19) | 562(22) | 670(15) | 345(14) | 698(24) | 1233(22) | 191(11) | 5519(19) |
| BMC, Barzilai Medical Center; BPMCP, Baruch Padeh Medical Center Poriya; BZMC, Bnei Zion Medical Center; HHUMC, Hadassah Hebrew University Medical Center; KMC, Kaplan Medical Center; MMC. Meir Medical center; RHCC, Rambam Health Care Campus; SMC, Shamir Medical Center; SZMC, Shaare Zedek Medical Center; TASMC, Tel Aviv Sourasky Medical Center; WMC, Wolfson Medical Center | | | | | | | | | | | | |
| * We included only healthcare workers who received the 3^rd^ vaccine dose between August 1^st^ 2021 and September 30^th^ 2021 | | | | | | | | | | | | |

**References:**

1. Israeli Ministry of Health COVID-19 data center. Accessed February 21, 2022. <https://data.gov.il/dataset/covid-19> (In Hebrew).
2. Patalon T, Gazit S, Pitzer VE, Prunas O, Warren JL, Weinberger DM. Odds of Testing Positive for SARS-CoV-2 Following Receipt of 3 vs 2 Doses of the BNT162b2 mRNA Vaccine. *JAMA Intern Med*. 2022; 182(2):179-184. doi:10.1001/jamainternmed.2021.7382
3. Bar-On YM, Goldberg Y, Mandel M, et al. Protection of BNT162b2 Vaccine Booster against Covid-19 in Israel. *N Engl J Med*. 2021; 385(15):1393-1400. doi:10.1056/NEJMoa2114255
